# Modelling the dispersion of SARS-CoV-2 on a dynamic network graph

**DOI:** 10.1101/2020.10.19.20215046

**Authors:** Patrick Bryant, Arne Elofsson

## Abstract

**Background:** When modelling the dispersion of an epidemic using R_0_, one only considers the average number of individuals each infected individual will infect. However, we know from extensive studies of social networks that there is significant variation in the number of connections and thus social contacts each individual has. Individuals with more social contacts are more likely to attract and spread infection. These individuals are likely the drivers of the epidemic, so-called superspreaders. When many superspreaders are immune, it becomes more difficult for the disease to spread, as the connectedness of the social network dramatically decreases. If one assumes all individuals being equally connected and thus as likely to spread disease as in a SIR model, this is not true.

**Methods:** To account for the impact of social network structure on epidemic development, we model the dispersion of SARS-CoV-2 on a dynamic preferential attachment graph which changes appearance proportional to observed mobility changes. We sample a serial interval distribution that determines the probability of dispersion for all infected nodes each day. We model the dispersion in different age groups using age-specific infection fatality rates. We vary the infection probabilities in different age groups and analyse the outcome.

**Results:** The impact of movement on network dynamics plays a crucial role in the spread of infections. We find that higher movement results in higher spread due to an increased probability of new connections being made within a social network. We show that saturation in the dispersion can be reached much earlier on a preferential attachment graph compared to spread on a random graph, which is more similar to estimations using R_0_.

**Conclusions:** We provide a novel method for modelling epidemics by using a dynamic network structure related to observed mobility changes. The social network structure plays a crucial role in epidemic development, something that is often overlooked.

## Introduction

Early this year, a new virus, SARS-CoV-2, emerged from China and quickly spread across the world. To limit the spread of the virus, governments introduced different strategies of suppression[1]. Extensive testing procedures were set up to follow the development of the epidemic and maintain control over the spread. To analyse the development of the spread accordingly, the basic reproductive number, R_0_, was estimated from confirmed cases using SIR models [1–3]. The objective of tracking R_0_ is to keep it below one, as this means a reduction in spread according to the SIR models and that the infection will disappear.

When modelling the spread of an epidemic using R_0_, one only considers the average number of individuals each infected individual will infect. However, we know from extensive studies of social networks[4–6] that there is variation in the number of connections and thus, social contacts each individual has. Individuals with more social contacts are more likely to attract disease. These individuals are also more likely to spread disease due to their many connections. These individuals are likely the early drivers of the epidemic, so-called super spreaders [7].

When the superspreaders are immunised; however, it becomes more difficult for the disease to spread, as the connectedness of the social network dramatically decreases. If one assumes all individuals are equally connected and thus as likely to spread disease as in the case of R_0_, this is not true [8]. To account for the tendency to spread to occur, one has to consider social network structure when modelling epidemic development.

Later in the epidemic, the large spread may therefore be unfavoured regardless of social contact, as there is much less opportunity for a virus such as SARS-CoV-2 to spread through the social network. This may explain why some countries (e.g. Sweden, Italy and the USA [9]) have increased mobility [10] due to lifting of restrictions, which is found to be correlated to spread [11], but continuously low numbers of cases and deaths.

To account for the impact of social network structure on epidemic development, we here model the dispersion of SARS-CoV-2 on a preferential attachment graph. We provide a novel method for modelling epidemics by sampling a serial interval distribution that determines the probability of spread for all infected nodes at each day. As compared to previous models utilising social networks, we don’t use R_0_ for spread [12]. An existing model that uses a SIR model in combination with a network does not assess their model’s accuracy in terms of observed cases or deaths, or the impact of the variable spread between different age groups [13]. A network model that used GPS data to estimate the impact of control strategies in the small town of Haslemere [14] used only 468 individuals and thereby only captured a minimal network. Here we provide a way to model spread in cities and countries of different sizes efficiently by scaling. We analyse the outcome of different simulations by comparing them to observed death tolls and estimate the impact of reducing spread in different age groups, a crucial aspect to consider due to the great variability in infection fatality rates between age groups [15,16].

## Methods

### Social network model

To model the dispersion of COVID-19, we approximate a social network by a preferential attachment model[17] with n nodes. In each step of the network generation, a node with m new links is introduced. The probability of existing nodes to obtain a link to the newly introduced one is:

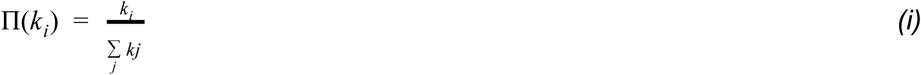

where ki is the degree of node i and ∑kj is the sum over the degree of all nodes n. The network is generated using “networkx” [18].

The network is made dynamic by rearranging the links between existing nodes according to changes in mobility patterns [10]. The number of links to be rearranged is chosen proportionally to the change in mobility pattern:

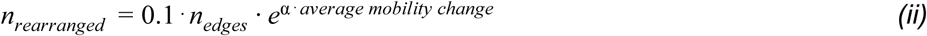

where n_edges_ are the remaining edges and the average mobility change is the average change across the six mobility sectors retail and recreation, groceries and pharmacies, transit stations, workplaces, residential and parks. The sign of the residential changes is reversed, as increased residential mobility should mean decreased probability of reconnecting links. Alpha is a scaling factor. If the average mobility change is 0, 10% of the remaining edges will thus be rearranged.

### Random network

To compare the spread on a preferential attachment network, which has a defined intrinsic structure, we model the spread on a random network [19]. To obtain a similar number of links as to the variations of m, we let p = 1/(n/2m). The network is also generated using networkx [18].

### Infection model

The infection model used is based on the classic SIR model [3]. We initialize the infection by a random uniform sampling of n_i_ nodes. The links of these nodes are then used to spread the infection. The probability that infection will spread from the sampled nodes is calculated through the serial interval[20] :

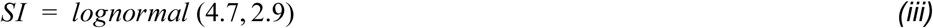

The serial interval is discretized in steps of 1 day:

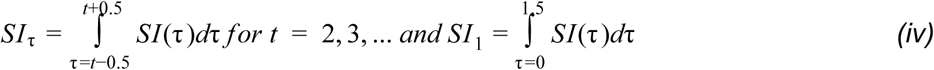

The number of infectious nodes at day τ, n_I,τ_, is thus:

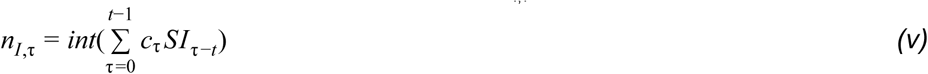

where c_τ_, are the number of cases (infections) at day τ.

The number of infectious nodes is rounded to the nearest integer and represents the number of nodes of the currently infected nodes that will spread the infection. The number of cases in each age group is modelled individually by dividing all n nodes into 4 age groups, according to the population shares in Tables 1 and 2. The number of infectious nodes at day τ, n_I,τ_ are randomly uniformly sampled from the infected nodes at day τ, which have not yet been selected to infect. For each of the nodes n_I,t,i_ in n_I,t_, all nodes which have a link to n_I,τ,i_, n_link,τ,i,ag_ in age group ag are selected to be infected. The total number of cases at day τ is thus:

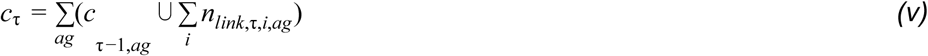

where ag = 1-4 are the age groups 0-19, 20-49, 50-69, and 70+. When a node has been selected to spread the infection to its connected nodes, it is removed and can thereby not spread any further infection.

**Table 1.** Infection fatality rates (IFRs) estimates for four different age groups in Spain [27] and population shares for the same age groups [28].

| Age | Population share (%) | IFR (%) |
| --- | --- | --- |
| 0-19 | 20 | 0.0 |
| 20-49 | 40 | 0.035 |
| 50-69 | 26 | 0.645 |
| 70+ | 14 | 5.8 |

**Table 2.** Infection fatality rates (IFRs) estimates for four different age groups in the USA [16] and population shares for the same age groups in New York City[29].

| Age | Population share (%) | IFR (%) |
| --- | --- | --- |
| 0-19 | 23 | 0.003 |
| 20-49 | 45 | 0.02 |
| 50-69 | 22 | 0.5 |
| 70+ | 10 | 5.4 |

To account for the fact that new infections can be introduced from outside of the already infected sub-network a pseudo count is added to c_τ_ at each day by randomly uniformly sampling the susceptible nodes (the nodes not infected or removed):

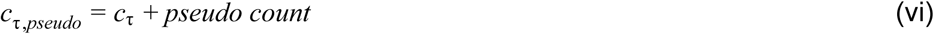

### Modifying the probability of spread

We assume the epidemic starts 4 weeks before at least 10 deaths have been observed. To account for the suppression methods, we introduce a reduction in the probability of a node selected to be infected individually for each age group proportionally to the change in mobility. The probability for infection per age group is varied by deciding if each node selected for the spread in that age group (see equation *(v)*) will indeed be infected by following a probability p_ag_. If the reduction in probability in an age group is to 25%, the nodes in that age group will only be infected in¼ cases.

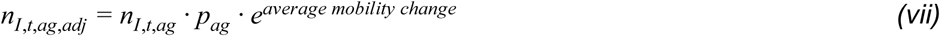

### Death model

To evaluate the model, we analyse the number of daily deaths in Spain [21] and New York City[22]. The number of daily deaths at day τ for age group ag, d_τ,ag_, is estimated per age group by using the number of cases per age group at day τ, c_τ,ag_, and the estimated infection fatality rates (IFRs) (see Tables 1 and 2):

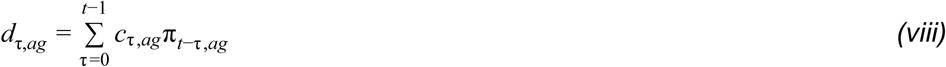

where π_τ,*ag*_ is the infection to death distribution for age group ag at day τ, a combination of the infection to onset distribution (Gamma(5.1,0.86)) and onset to death distribution (Gamma(17.8,0.45)) (combined with mean 22.9 days and standard deviation 0.45 days) times the infection fatality rate (IFR) [23],[24],[25] :

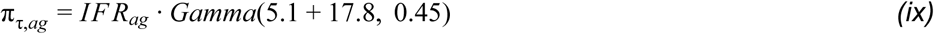

Π_t,ag_ is discretized in steps of 1 day accordingly:

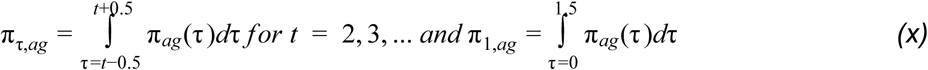

To simulate the dispersion in a network of large size, we scale the number of modelled daily deaths with the population size divided by the number of nodes in the network (n=10000).

The population size for Spain is X and for New York City[26]. The number of daily adjusted deaths for age group ag at day τ are thus:

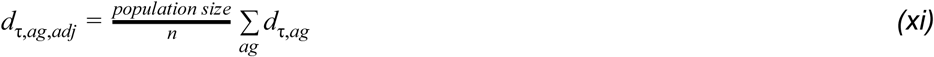

### Varying the number introduced links (m) and the infection probability

To investigate the effect of network structure (m) we set n to be n1=10000 and m to be 1 and 2. The probability of a node to be infected if linked to an infectious node (see *equation (vii)*)is varied as 50, 33 and 25% for all the age groups. This probability is kept constant throughout the simulation. The spread is thus only influenced by network dynamics and mobility changes. The number of initial nodes for n1 is 1 and for n2 10, both with matching pseudo counts of 1 and 10 respectively (see *equation (vi)*).

### Seed initialization for preferential attachment networks

To adjust for the randomness in network generation, we initialize all networks of different m with 3 different random seeds. The average results from the 3 simulations are then compared with each other.

### Code

The code is freely available under the GPLv3 license. https://github.com/patrickbryant1/epidemic_net

## Results

### Deaths

When analyzing the number of daily deaths and the modelled outcomes, it is clear that m=1 and 33% reduction in infection probability gives the best fits for Spain and m=2 and 33% reduction in infection probability for New York City, using 10000 nodes (Figures 1 and 2). The outcomes from the different combinations of infection probabilities vary widely for different m in both Spain and New York, while the alphas seem to have little impact. The higher m is, the more deaths are observed, regardless of the infection probabilities. When m=2, all combinations of infection probabilities give high death tolls.

**Figure 1.**
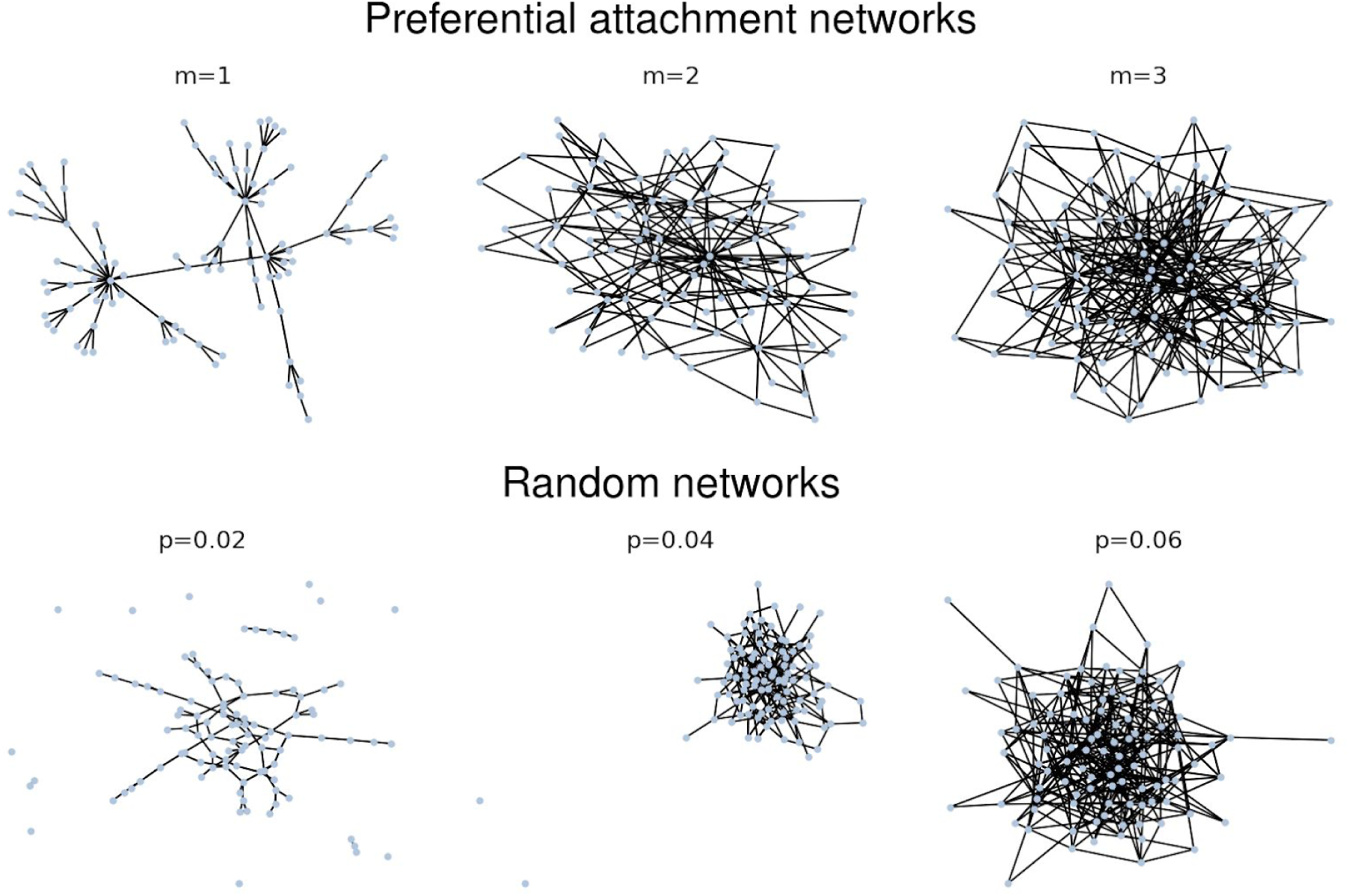
Visualization of preferential attachment and random networks for different m and corresponding p, using 100 nodes.

**Figure 1.**
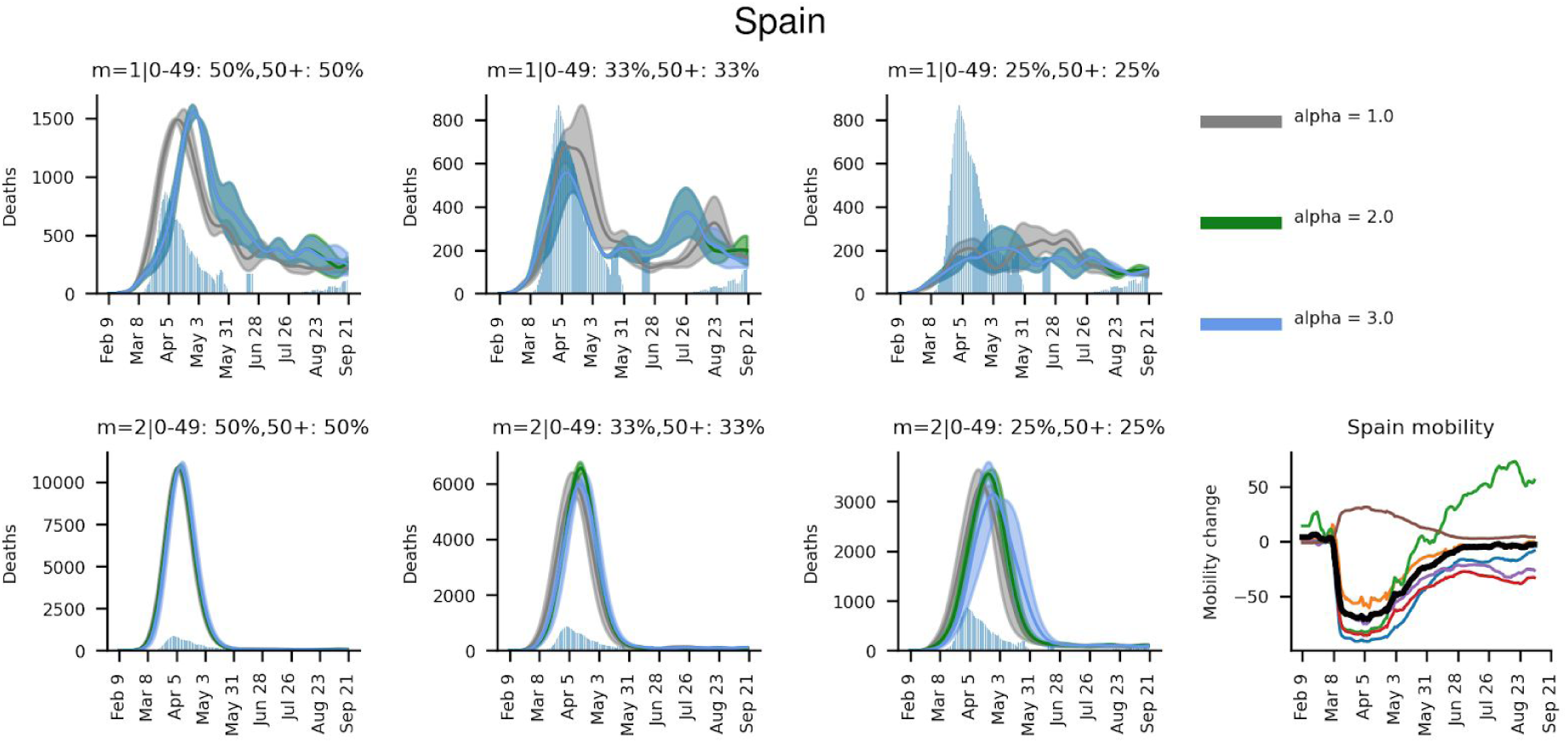
Daily deaths (averages from 3 simulations) for Spain using a preferential attachment network graph with 10000 nodes. The grey histogram represents the observed deaths, smoothed using a 7-day window. The number of links introduced with each new node, m, is varied from 1-3 and the infection probabilities in the age groups 0-49 and 50+ are varied in 3 different combinations, ranging from 50% reduction to 25%. The impact of the mobility change on the network dynamics, alpha, is varied from 1 to 3. Note that the deaths are scaled with a factor according to *equation (xi)*.

**Figure 2.**
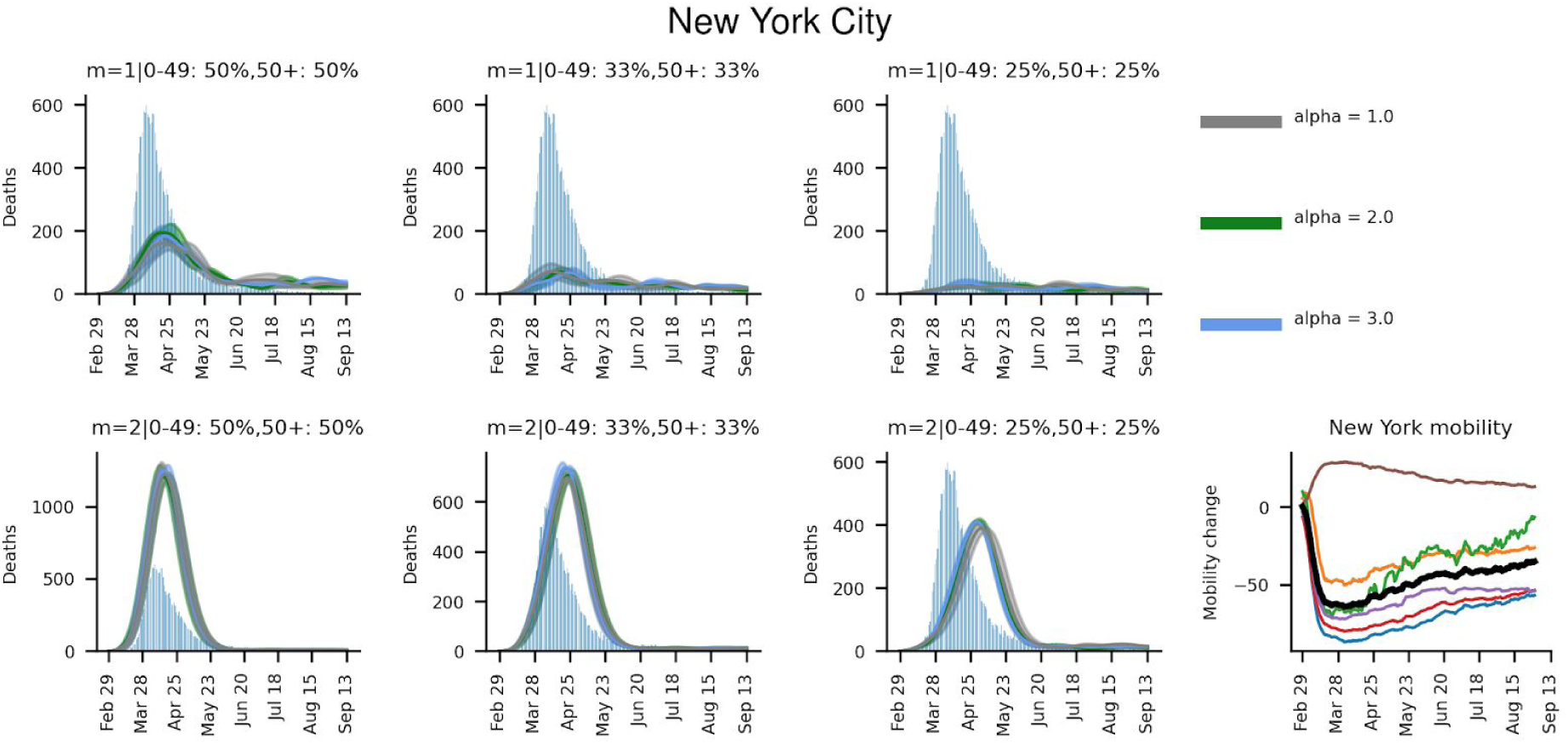
Daily deaths (averages from 3 simulations) for NewYork City using a preferential attachment network graph with 10000 nodes. The grey histogram represents the observed deaths, smoothed using a 7-day window. The number of links introduced with each new node, m, is varied from 1-3 and the infection probabilities in the age groups 0-49 and 50+ are varied in 3 different combinations, ranging from 50% reduction to 25%. The impact of the mobility change on the network dynamics, alpha, is varied from 1 to 3. Note that the deaths are scaled with a factor according to *equation (xi)*.

The relationship between the daily number of deaths and the infection probabilities and alphas is not at all straightforward. Considering m=1 for both Spain and New York City, widely different developments in the daily deaths can be observed (Figures 1 and 2 respectively). Spain, which has increased its mobility back to 0, observed a resurgence in the spread. New York City, which has kept the mobility low does not. This is true regardless of what alpha is used.

Compared to spread on a random network, both Spain and New York City observe faster spread and saturation with the preferential attachment model. In New York City, for m=1 and 50% reduction in infection probability, the peak deaths are observed in middle of April on the preferential attachment graph, while the peak is in middle of June on the random network (Figures 2 and S2 respectively). The same is true for Spain, peak in beginning to middle May on a random graph compared to beginning to middle of April on a preferential attachment graph, for m=1 and 50% reduction in infection probability (Figures S1 and 1 respectively).

### Cases

Almost identical levels of saturation are observed in both Spain and New York City, despite the fact that both their model parameters and mobility changes differ greatly. In contrast to the herd immunity estimates obtained when using the basic reproductive number [30] (around 80% for Spain and 70% for the USA), the level of saturation is found to vary greatly between a few per cent for m=1 to 80% for m=2 (Figures 3 and 4). As expected, the least connected network (m=1) yields the lowest saturation point, while the most connected one (m=2) yields the highest. In the case of m=1, the spread is not saturated at the end of the simulations, regardless of infection probability. For m=2, the saturation occurs at around 30-80%. The case that most closely resembles New York City (m=2, 0-49:33%, 50+:33%) saturates at 50%, while the best fit for Spain (m=1, 0-49:33%, 50+:33%) indicates continued, but slowing spread at around 12.5%.

**Figure 3.**
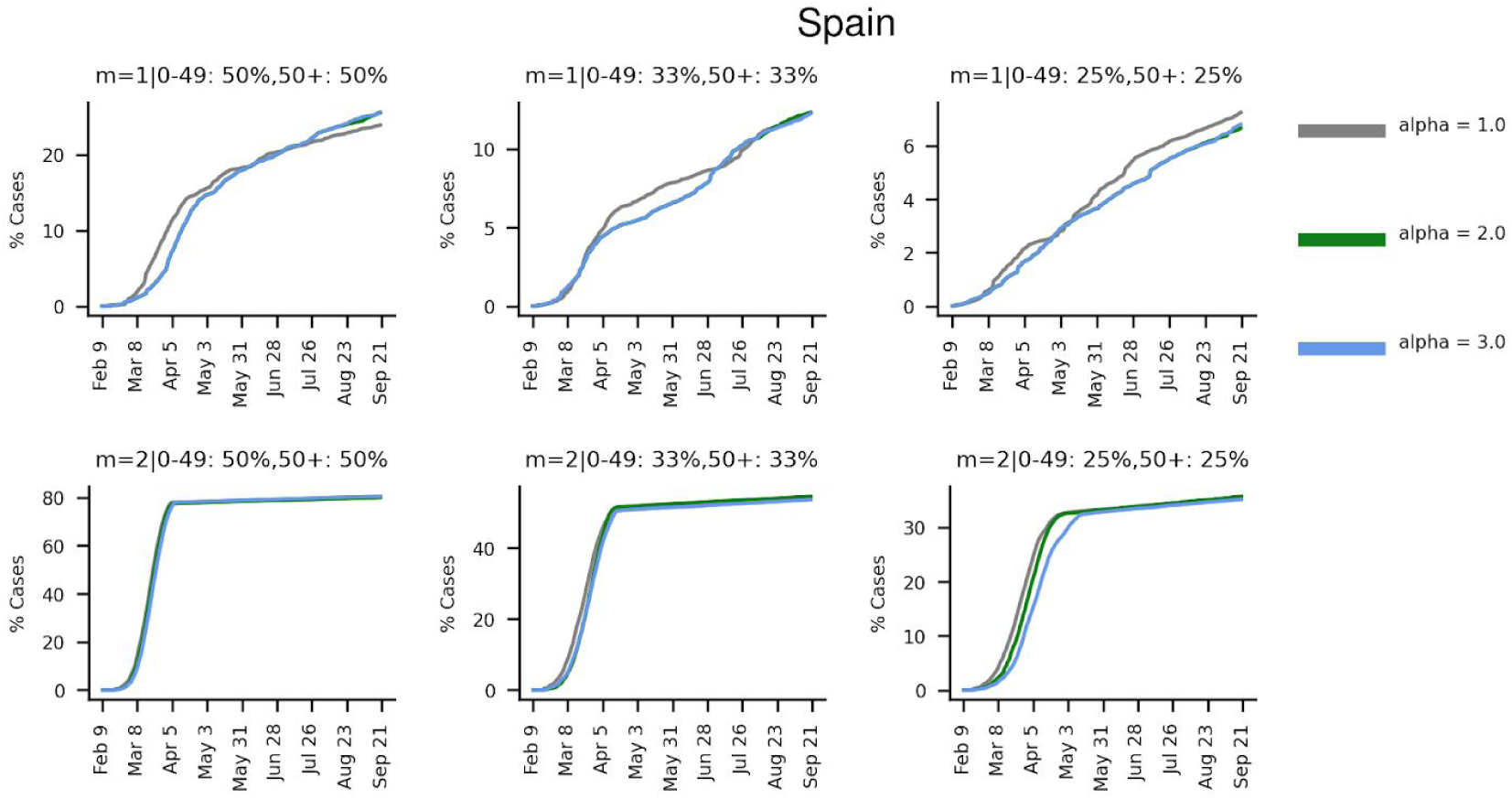
Cumulative% cases of the total number of nodes (n=10000) for Spain (averages from 3 simulations). The solid lines represent the average modelled outcomes, while the dashed lines represent the standard deviation. The number of links introduced with each new node, m, is varied from 1-3 and the infection probabilities in the age groups 0-49 and 50+ are varied in 3 different combinations, ranging from 50% reduction to 25%. The impact of the mobility change on the network dynamics, alpha, is varied from 1 to 3.

**Figure 4.**
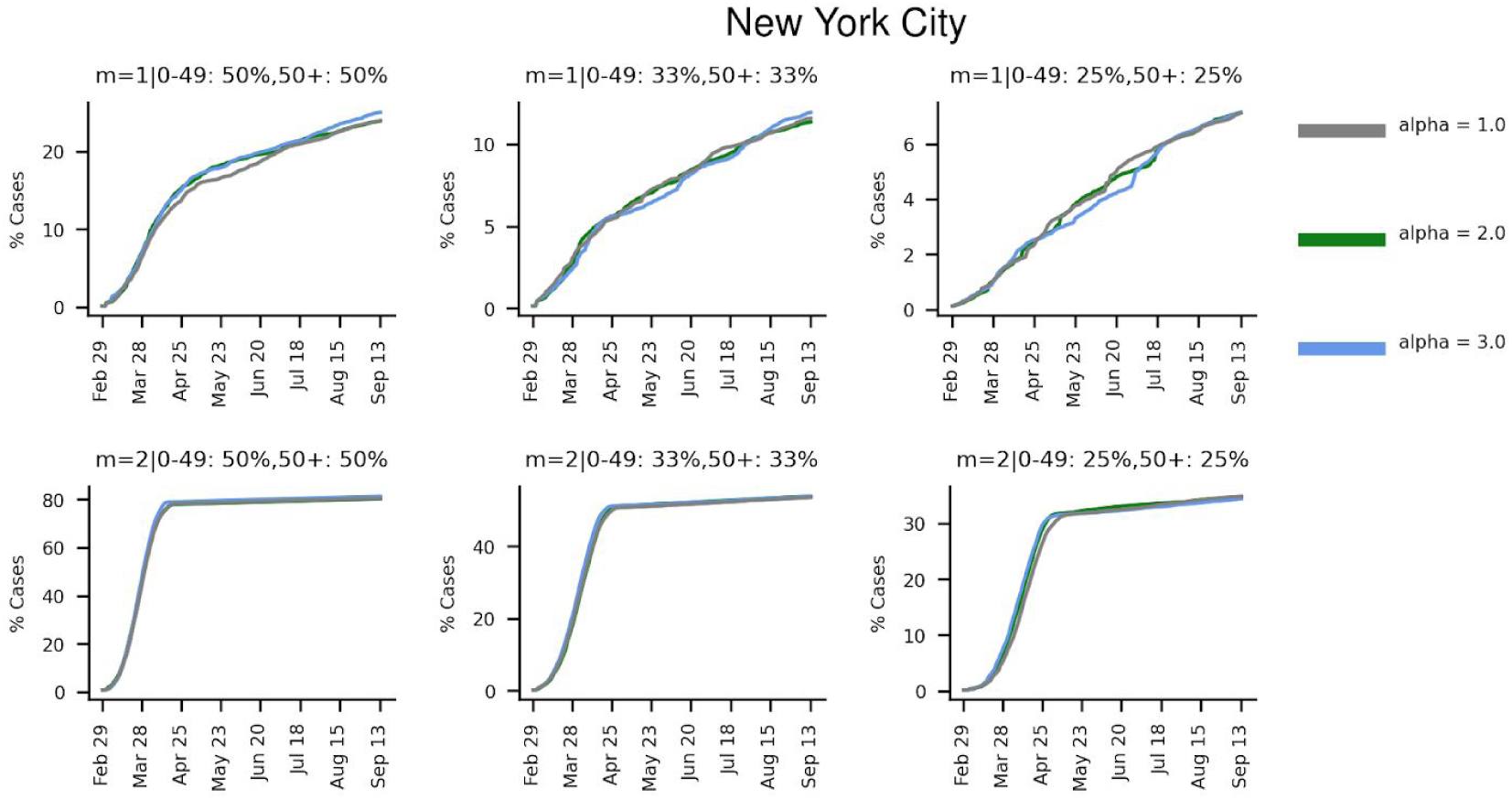
Cumulative% cases of the total number of nodes (n=10000) for New York City (averages from 3 simulations). The solid lines represent the average modelled outcomes, while the dashed lines represent the standard deviation. The number of links introduced with each new node, m, is varied from 1-3 and the infection probabilities in the age groups 0-49 and 50+ are varied in 3 different combinations, ranging from 50% reduction to 25%. The impact of the mobility change on the network dynamics, alpha, is varied from 1 to 3.

### Node degree and removal

The proportion of nodes with a degree above threshold t (t=5*average degree) gives a measure of how connected the network is (Figures 5 and 6). If this proportion saturates quickly, the network’s most connected nodes are retained, which means the network can become disconnected very easily. As can be seen for all m, the infection probability combinations that result in the highest death tolls are also those that are among the least efficient in disconnecting the networks.

**Figure 5.**
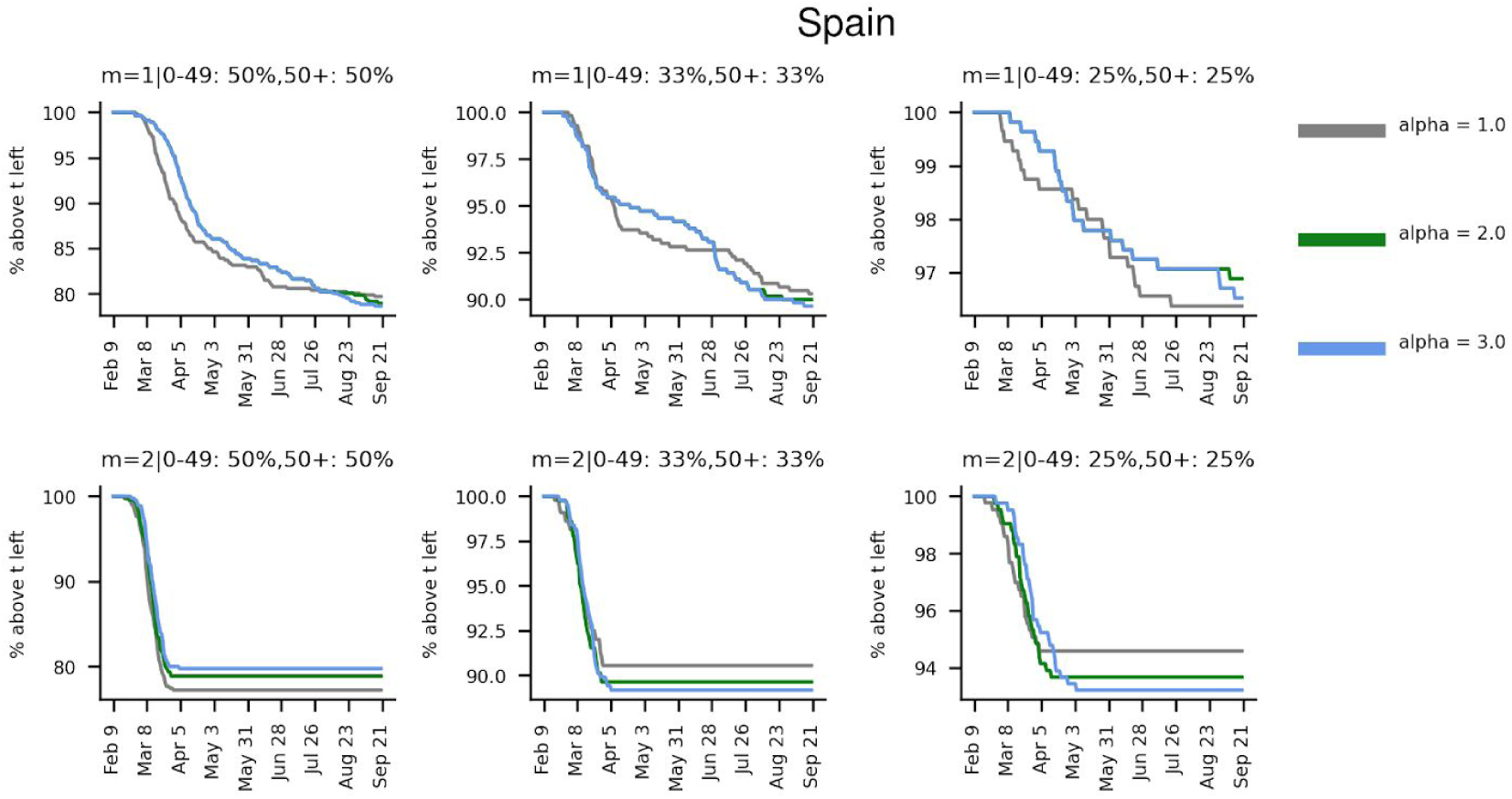
The fraction of nodes that are not infectious above threshold t (t=5·average degree, n=10000) for Spain (averages from 3 simulations). The solid lines represent the average modelled outcomes, while the dashed lines represent the standard deviation. The number of links introduced with each new node, m, is varied from 1-3 and the infection probabilities in the age groups 0-49 and 50+ are varied in 3 different combinations, ranging from 50% reduction to 25%. The impact of the mobility change on the network dynamics, alpha, is varied from 1 to 3.

**Figure 6.**
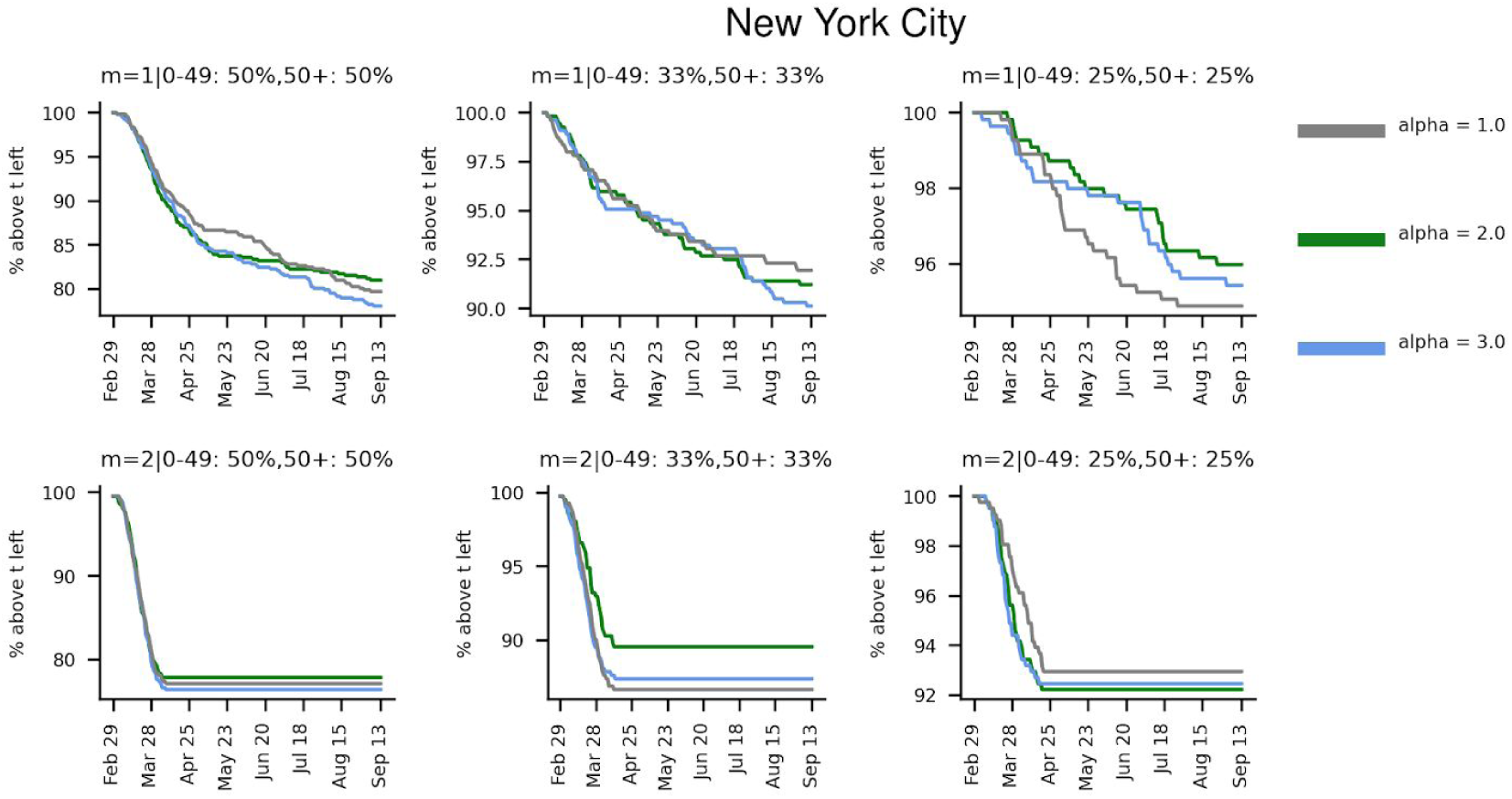
The fraction of nodes that are not infectious above threshold t (t=5·average degree, n=10000) for New York City (averages from 3 simulations). The solid lines represent the average modelled outcomes, while the dashed lines represent the standard deviation. The number of links introduced with each new node, m, is varied from 1-3 and the infection probabilities in the age groups 0-49 and 50+ are varied in 3 different combinations, ranging from 50% reduction to 25%. The impact of the mobility change on the network dynamics, alpha, is varied from 1 to 3.

For both Spain and New York City, the saturation points for m=1 are not reached for any infection probabilities at the end of the modelling. As of September 21 in Spain and September 13 in New York City, the remaining fraction of nodes that are not infectious above t (5*average degrees) are 80, 90 and 97% and 80, 90 and 96% for the infection probabilities 50, 33 and 25% respectively.

## Discussion

To account for the variation in the number of connections each individual has, epidemic development has to be modelled on a network graph. Modelling the spread of an epidemic using the basic reproductive number, R_0_, as in a classical SIR model [3], one does not account for the effect of social network structures [7] on the spread. Further, a real social network will be dynamic and dependent on movement. The preferential attachment graph network we use in our model provides an opportunity to assess different scenarios of dispersion on different dynamic network structures and to analyze the importance of varying infection probabilities and mobility impact in different age groups.

Since there is no available data on the structure of the social networks or the interactions between different age groups in Spain or New York City, we model different scenarios, varying m from 1 to 3 with 3 different combinations of infection probabilities. We make the network dynamic according to observed changes in mobility patterns and let these changes impact the network dynamics scaled by a factor, alpha, which we vary in three different combinations. Even though the value of alpha seems to have little impact on the spread, the mobility changes appear to influence the outcome substantially. Compared to spread on a random network, both Spain and New York City observe faster spread and saturation with the preferential attachment model due to the structural differences of the two network types.

The mobility data indicates that movement is back to normal levels in Spain, while it is still low in New York City. Further, the case simulations show saturation in New York City, while the spread is still ongoing in Spain. New York City is denser and likely more connected than Spain, having a population density of 10400 inhabitants per km^2^ (data from the year 2010, [31]) vs 94 in Spain (data from the year 2018, [32]). This is consistent with m=1 giving the best fit for Spain, but m=2 for New York City. New York City should thereby have a higher level of saturation in terms of the number of cases. The precision in New York City is much greater compared to modelling a whole country like Spain, which the biggest and most connected cities likely contribute to most of the observed spread.

When having a highly connected network, there is a higher probability for the spread of the infection to continue in an alternative way, even if the infection probability is reduced, as can be seen in the higher case percentages in Figures 3 and 4 for higher m. In the case of m=2 the cumulative cases are therefore saturated quickly due to high spread. In contrast, for m=1 the spread is not saturated at the end of the simulations, displaying a slower, more stable spread. The modelled infection level of 50% in New York City is higher than the observed levels of around 30% (https://www.nytimes.com/2020/08/19/nyregion/new-york-city-antibody-test.html, last accessed Sep 16 2020). The modelled infection level for Spain is around 5% at the beginning of May corresponding well with the reported level (5% for at April 27-May 11 [33]).

The faster a network is disconnected, and the more highly connected nodes are retained at that point, the less dispersion is likely to have happened in that network. This means that even if infection probabilities are low, more spread can happen if many highly connected nodes have to be removed before the spread in the network is saturated. In the modelling of New York City (Figure 6), the spread is saturated while 90% of the nodes that are not infectious above degree threshold t (5·average degree) remain for m=2 and infection probability 33%, while for m=1 and infection probability 33%, the spread continues at 90%. This peculiarity arises due to the m=2 being much more connected and infecting 10% of the highly connected nodes there will result in a much higher spread than doing the same with m=1.

Modelling spread on networks generated from random processes is not optimal. We, therefore, generate three networks for each combination of m and infection probabilities. The outcome will still be dependent on which nodes are picked to disperse the infection at what time and how they are connected. The outcomes are dependent on the number of initial nodes, the percentage of nodes that are reconnected at each step and the pseudo counts as well. The necessity for scaling the results due to limitations in computational resources can also influence the results. Optimally, the dispersion should be modelled on a known social network, where both the number of initially infected individuals, their movements and their contacts are known. In lack of such data, we provide the best available approximation of scaled dynamic preferential attachment graphs with varying structure, infection probabilities and observed mobility changes.

## Conclusions

When a network is more connected, both faster and more spread is observed. The reduction in infection probability matters, but is secondary to the network structure. The impact of the mobility change on spread displays no straightforward relationship since the parameter alpha has no substantial impact on the outcome. The mobility seems to impact the outcome though, since the modelling of New York city displays no second peak, keeping the mobility low, while Spain increased its mobility levels to normal and displays a resurgence in the spread. The relationship between spread, network structure, infection probability and network dynamics is complex and needs to be modelled at great detail to obtain reliable estimates of spread, something that has proven difficult to do with available data.

## Data Availability

The code is freely available under the GPLv3 license.

https://github.com/patrickbryant1/epidemic_net

## Declaration of interests

We declare no competing interests.

## Funding statement

### Financial support

Swedish Research Council for Natural Science, grant No. VR-2016-06301 and Swedish E-science Research Center. Computational resources: Swedish National Infrastructure for Computing, grant No. SNIC-2019/3-319.

## Supplementary material

**Figure S1.**
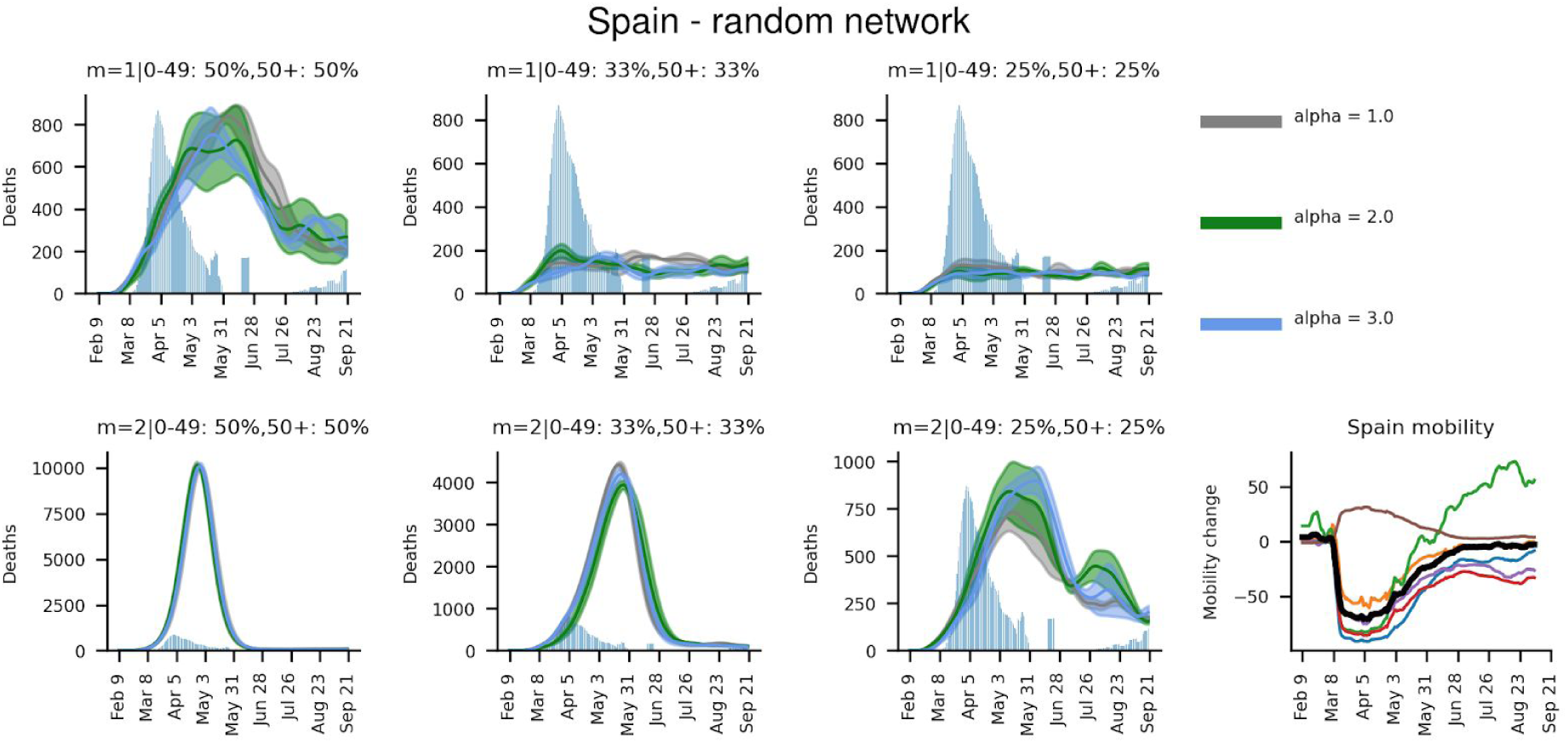
Daily deaths (averages from 3 simulations) for Spain using a random network graph with 10000 nodes. The grey histogram represents the observed deaths, smoothed using a 7-day window. The number of links introduced with each new node, m, is varied from 1-3 and the infection probabilities in the age groups 0-49 and 50+ are varied in 3 different combinations, ranging from 50% reduction to 25%. The impact of the mobility change on the network dynamics, alpha, is varied from 1 to 3. Note that the deaths are scaled with a factor according to *equation (xi)*.

**Figure S2.**
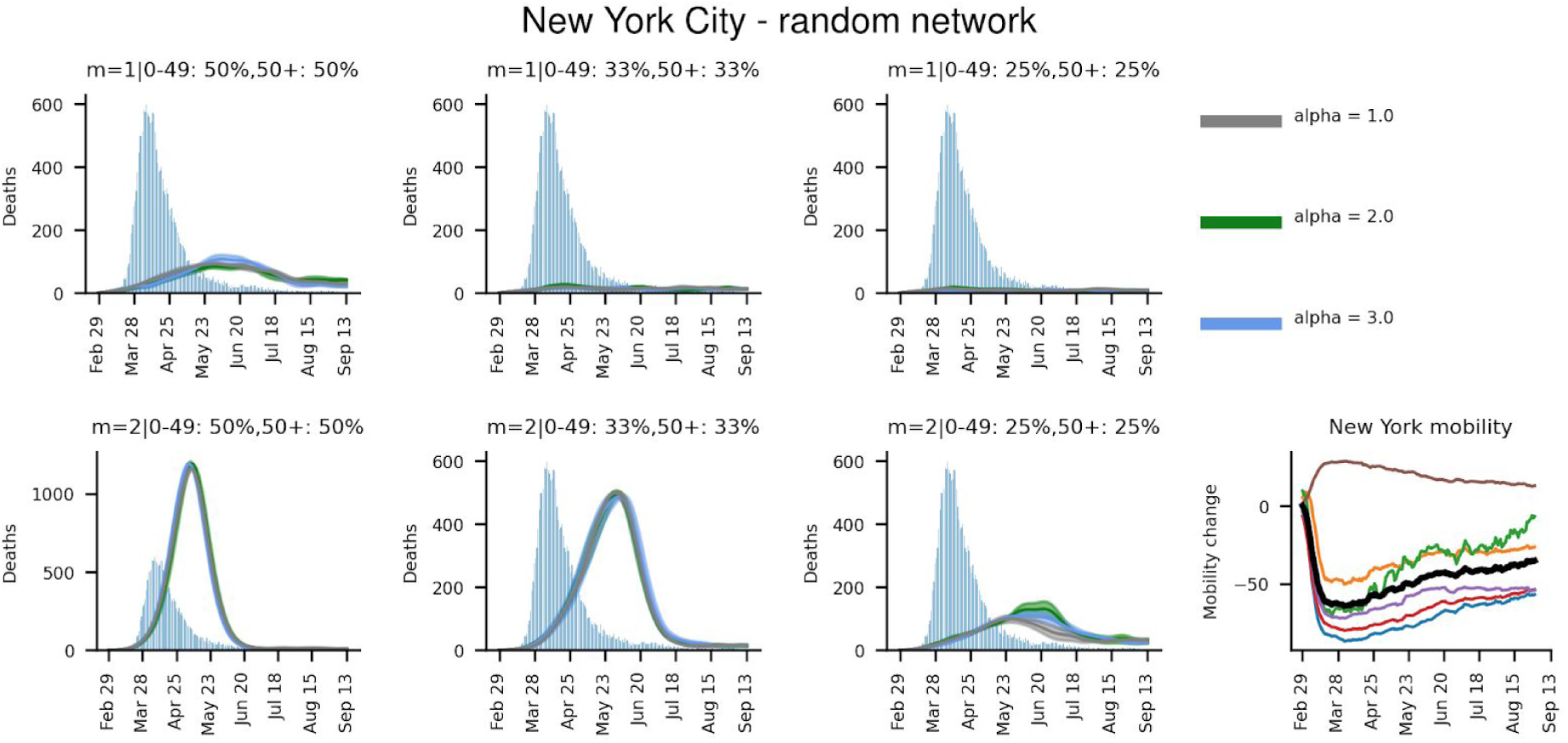
Daily deaths (averages from 3 simulations) for New York City using a random network graph with 10000 nodes. The grey histogram represents the observed deaths, smoothed using a 7-day window. The number of links introduced with each new node, m, is varied from 1-3 and the infection probabilities in the age groups 0-49 and 50+ are varied in 3 different combinations, ranging from 50% reduction to 25%. The impact of the mobility change on the network dynamics, alpha, is varied from 1 to 3. Note that the deaths are scaled with a factor according to *equation (xi)*.

## Notes

### Competing Interest Statement

The authors have declared no competing interest.

